# COVID-19 Beliefs, Behaviors, and Perceptions Among Students on a College Campus During the Global Coronavirus Pandemic

**DOI:** 10.1101/2024.12.23.24319584

**Authors:** Avian White, Guy Iverson, LaNika Wright, John T. Fallon, Charles Humphrey

## Abstract

**Objectives:** Elucidating how people think and behave during a disease outbreak may provide valuable insight and help direct programs or surveillance to combat the spread of disease. As universities welcomed students back to their campuses following COVID-19 shutdowns, it became important to know students’ beliefs on COVID-19 and how these beliefs guided behaviors?

The objective of this study was to determine how students at East Carolina University felt about COVID-19, which behaviors they exhibited during the pandemic, and whether their feelings and behaviors differed significantly between the Spring and Fall 2021 semesters.

**Methods:** Surveys (N= 408) were distributed to students who were currently enrolled in Environmental Health Science classes during the Spring and Fall semesters of 2021. Questions were developed using a Likert scale and were analyzed to determine significant differences (p < 0.05) between semesters. Statistical analyses were performed using SPSS (SPSS Institute, Chicago, Ill).

**Results:** Results showed most students felt “somewhat concerned” about the COVID-19 pandemic during both semesters. Significant differences in student concern regarding COVID-19 between Spring and Fall semesters, were not observed *p* = 0.598. Student behaviors regarding weekly gatherings significant differed between semesters with a reported increase in gatherings of 5+ during the Fall semester, *p* < 0.001. Interestingly, we found more students indicated during the Spring semester in comparison to the Fall that they believed the vaccine was not safe and they would not take it (*p* < 0.001).

**Conclusion:** Our findings suggest that as the pandemic went on, behavior changes were observed in students between the semesters. This information may be important to officials as cases may fluctuate over time. Knowledge of attitudes and/or behaviors and awareness may help explain these fluctuations, allowing public health professionals to adjust recommendations and focus more intently on populations at risk.

## Introduction

Surveys can be a useful tool in gathering population information if they are properly developed, administered, and analyzed. Surveys with questions that solicit information such as private behaviors, mental health, and attitudes from persons may result in more reliable and valid data.^1,2^ Generally, surveys should be administered to a representative sample of the population of interest, thus allowing for generalization of results to the entire population.^3,4,5^ This collection of information may be of particular importance during times of disease outbreaks. Because infectious diseases have the capacity to rapidly expand across populations, the ability to collect efficacious data quickly may have a profound effect on public health policy development.^6^ The ease in which some surveys can be delivered may provide a cost-effective means of gathering necessary data.^7^ Additionally, responses from surveys may help public health agencies determine populations most at risk, as used during the height of the HIV/AIDS epidemic. This may help when determining and rolling out prevention and disease mitigation programs. Thus, surveys have been useful in healthcare epidemiology.

Recently, surveys have been used to gather information from specific populations during the COVID-19 outbreak.^8,9,10^ One such study in Guangdong, Province in China sought to determine the mental effects and awareness in college students regarding the disease and their association with future health behaviors. Researchers found older students had greater levels of awareness and higher-level changes in future behaviors.^11^ Other surveys have shown an increased prevalence of psychological health problems and a negative association with COVID-19 awareness levels in students.^12^ Increased levels of disease awareness may have an impact on behaviors that could contribute to the spread of disease and affect public health.^13,14,15^ Research conducted on COVID-19 awareness has shown that those with increased awareness were more likely to adopt strategies such as mask-wearing, social distancing, and handwashing.^16,17,18,19^ This suggests that information gleaned from these surveys may be used to develop specific cost-effective prevention strategies for targeted populations based on their needs.^20,21^

Students on the campus of ECU were surveyed to determine their beliefs, attitudes, and practices during the COVID-19 outbreak. It was hypothesized that there would be significant differences with student attitudes and beliefs concerning COVID-19 between the Spring and Fall semesters, with students becoming less concerned as vaccines become available and pandemic fatigue grew. Additionally, we sought to determine whether behavioral changes occurred between semesters with students becoming more relaxed with their manners (i.e., handwashing, congregating in large groups, etc.) after the rollout of vaccines.

## Methods

### Participants

Surveys were distributed to students who were currently enrolled in Environmental Health Science classes during the Spring and Fall semesters of 2021. Environmental Health Science 2110 qualifies as is a natural science, general education requirement, thus students may be representative of any undergraduate degree program at ECU since each student must complete these foundational requirements. from multiple majors (e.g., psychology, pre-health, elementary education, etc.) enroll in this class. Students had to be at least 18 years of age and actively enrolled to participate in the survey. Student participants in these classes ranged from freshmen to seniors. Additionally, master’s students enrolled in Environmental Health Science 6210, a graduate writing series class, were also invited to complete the survey. Data were collected from students using an online anonymous survey created in and distributed through Research Electronic Data Capture (RED Cap) survey system hosted at ECU.

### Ethics Statement

This project was reviewed the East Carolina University and Medical Center Institutional Review Board and was classified as exempt as it did not collect personal identifiers (UMCIRB# 21-00186). A formal statement of consent was obtained via the through Research Electronic Data Capture (RED Cap) survey system prior to starting the survey.

### Survey

The survey included demographic questions regarding gender, race and ethnicity in addition to information concerning housing status (e.g., on- or off-campus) and education levels. Demographic data were compiled from the annual census completed by university administration in Fall 2021 to compare the sample population to the broader student population. To determine student beliefs and behaviors, participants were asked questions about virus perceptions, hand washing and mask wearing. Additionally, students answered questions about their social and learning experiences during the semester. Students in the Fall semester were given an additional question concerning the impact of university transitions from different class types (e.g., online to face-to-face) (Supplemental figure 1).

### Statistical Analysis

Cross tabulations were performed to compare responses by different groups. Questions were developed using a Likert scale and were analyzed to determine significant differences (p < 0.05) between the Spring and Fall 2021 semesters. For nonparametric data, a Kruskal-Wallis test was used to determine significance between data consisting of more than 2 samples, while Mann-Whitney testing was used to analyze significance between 2 samples. Statistical analyses were performed using SPSS (SPSS Institute, Chicago, Ill). SPSS analysis also included a post-hoc pairwise Mann-Whitney with Bonferroni adjustments.

## Results

Surveys (n = 408) were disseminated to students enrolled in environmental health classes during the Spring 2021 semester, with 102 responses received (25.2% response rate). During the Fall, 243 surveys were administered with 99 responses received (40.7% response rate). The sample population for the surveys exhibited similar racial percentages as the greater ECU student population (Table 1). More specifically, the racial percentage of surveyed students were within 5% of the general population for the two largest groups (African American and Caucasian). Males and Hispanic ethnicity were both underrepresented in the survey relative to the broader student population. The percentage of female respondents were approximately 17% greater than the percentage of females at ECU, while the percentage of male respondents was approximately 19% lower than the percentage of ECU’s general population that identifies as male.

**Table 1.** Demographic data of student respondents and the Fall 2021 university census. Demographic data for survey respondents were summarized by semester and pooled. Asterisks (*) indicate data was not available.

|  |  | Spring 2021 | Fall 2021 | Overall | ECU<br>Fall<br>2021 |
| --- | --- | --- | --- | --- | --- |
| **Gender** |  | *n* (%) | *n* (%) | % | % |
|  | Male | 22 (21.6) | 23 (23.2) | 22.4 | 41 |
|  | Female | 79 (77.5) | 74 (74.7) | 76.1 | 59 |
|  | Prefer not<br>to answer | 1 (1) | 2 (2) | 1.5 | * |
| **Race** |  |  |  |  |  |
|  | African-American | 15 (14.7) | 17 (17.2) | 16 | 16.7 |
|  | Caucasian | 66 (64.7) | 68 (68.7) | 66.7 | 63.8 |
|  | Asian | 8 (7.8) | 3 (3) | 5.5 | 2.8 |
|  | Native<br>American | 0 (0) | 1 (1) | * | * |
|  | 2 or More | 6 (5.9) | 1 (1) | * | * |
|  | Other | 7 (6.9) | 9 (9.1) | 8.5 | 8.7 |
| **Ethnicity** |  |  |  |  |  |
|  | Hispanic or<br>Latino | 6 (5.9) | 15 (15.2) | 10.5 | 8 |
|  | Not<br>Hispanic or<br>Latino | 96 (94.1) | 84 (84.8) | 89.5 | 92 |

### Student Attitudes and Beliefs Concerning COVID-19

Overall, it was shown that many students were somewhat concerned (46.5%) about the pandemic and believed it to be very transmissible (59.2%). Further analysis showed there was no significant difference in those feeling somewhat concerned between the Spring and Fall semesters, X^2^ (3, N= 200) = 1.877, *p* = 0.598. The number of those who felt the virus was “very transmissible” slightly decreased from Spring (5.9%) to Fall (5.1%) semesters, however this difference was not statistically significant X^2^ (2, N= 201) = 0.921, *p*= 0.631. Interestingly, 5.5% of the population felt the virus was very transmissible but were not concerned at all about the pandemic. It is possible these persons felt that while transmissible, the virus did not cause significant sickness and thus were not concerned.

While most classes during the Spring semester 2021 were online, dorms were open at limited capacity and students had the option to live in dormitories. Of the students who planned to live on campus, most (67.8%) felt safe with the housing arrangements. When analyzed by semester, 61.5% of persons felt safe on campus during the Spring semester with the number rising to 72.7% for the Fall semester. This perception of safety may have affected a student’s decision to stay on campus, in addition to capacity limitations. When students who lived off campus were queried, 31.6% said the COVID-19 pandemic affected their campus stay during the Spring semester, while 47.8% of those during the Fall semester said the pandemic affected their stay. It is also possible that online classes and the ability to go to school from home may have contributed to the Spring semester having a lower percentage of students on campus being affected by COVID-19.

Overall, there was some concern about COVID-19 vaccine with only 45.8% responding they felt “very safe taking it” and 69.7% felt the vaccine was at least a little safe and would take it (Table 2). These concerns may have impacted the overall student vaccination rate, 74%, observed by ECU by the end of the fall semester (During the Spring semester, prior to full roll out of the vaccines, 30% of the student population felt the vaccine was “not safe and would not take it”, while only 7.1% of students felt this way during the Fall semester. There was an almost 23% increase in the percentage of students surveyed that perceived the vaccine as being safe from Spring to Fall. Chi-square analysis showed significant differences between the semesters X^2^ (3, N= 201) = 18.418, *p* < 0.001. For those who did not feel safe taking the vaccine, the predominant reason was “untrusting of medical professionals”. Others noted “fear of long-term side effects in a vaccine that had a very short study time” and “too politicized” as reasons for not wanting the vaccine.

**Table 2.** Responses to questions regarding vaccine safety stratified by race and pooled between the Spring and Fall 2021 semesters.

|  | Yes, very<br>safe taking<br>it | A little safe,<br>but would<br>still take it | Not safe, but<br>would still<br>take it | Not safe, and<br>would not<br>take it |  |
| --- | --- | --- | --- | --- | --- |
| Race |  |  |  |  | Total |
| African American | 14 (15.2) | 8 (16.7) | 5 (21.7) | 5 (13.2) | 32 (15.9) |
| Caucasian | 60 (65.2) | 33 (68.8) | 16 (69.6) | 25 (65.8) | 134<br>(66.7) |
| Asian | 7 (7.6) | 2 (4.2) | 1 (4.3) | 1 (2.6) | 11 (5.5) |
| Native American | 0 | 1 (2.1) | 0 | 0 | 1 (0.5) |
| 2 or More | 3 (3.3) | 2 (4.2) | 0 | 2 (5.3) | 7 (3.5) |
| Other | 8 (8.7) | 2 (4.2) | 1 (4.3) | 5 (13.2) | 16 (8.0) |
| Total | 92 | 48 | 23 | 38 |  |

When reviewing vaccine perception data, those feeling very safe taking the vaccine did not differ much based-on gender. However, slightly more males, 24% responded as “not safe and would not take it” compared to 17% of females, a 7% difference. Overall, the percentage of students who would take the vaccine was 81.1%, regardless of the varying feelings of safety.

### Student Behaviors During Spring and Fall 2021 Semesters

To help mitigate the spread of COVID-19 among persons, the CDC gave advice on preventative behaviors such as hand washing, visiting, and mask wearing. The CDC promoted hand hygiene as a mitigation measure for COVID-19.^22^ Observations of student responses in this survey showed frequency in daily handwashing behavior was almost evenly split between 4-6 times (24.9%), 7-8 times (21.8%), and 9+ times (22.7%). Additionally, the pandemic may have affected how students visited others or allowed visitors. Overall, 39.0% of students reported having visitors at least 1-2 times during the week, 28.5% of respondents reported not having any visitors. To help reduce the likelihood of spreading the virus, off campus students were discouraged from visiting dorms and students were cautioned against congregating in large groups whether on or off campus during the Spring semester. Significant differences were observed between the semesters (*p*= 0.005). Notably, an increase in visitor frequency during the Fall semester and less students reported not having visitors during the Fall was observed among respondents.

Although mask wearing was strongly encouraged, only 5.7% reported always wearing a mask when having visitors and 39.9% reported never wearing a mask. A higher percentage of students reported wearing masks during the Spring relative to Fall semesters, and the differences were statistically significant (*p* = 0.042). When analyzing mask wearing between genders, we found no statistical difference (*p* = 0.161). It’s possible that COVID perceptions may have influenced mask wearing when campus students were entertaining visitors. Visitation may have also been influenced by the type of visitor (e.g., family, close friend), however we were not able to gather that information from this survey. Chi-square testing showed a significantly significant (*p* = 0.001) association between transmissibility perception and mask wearing. Notably, almost all persons who perceived the virus as not very transmissible, reported never wearing a mask (87.5%) (Figure 1).

**Figure 1.**
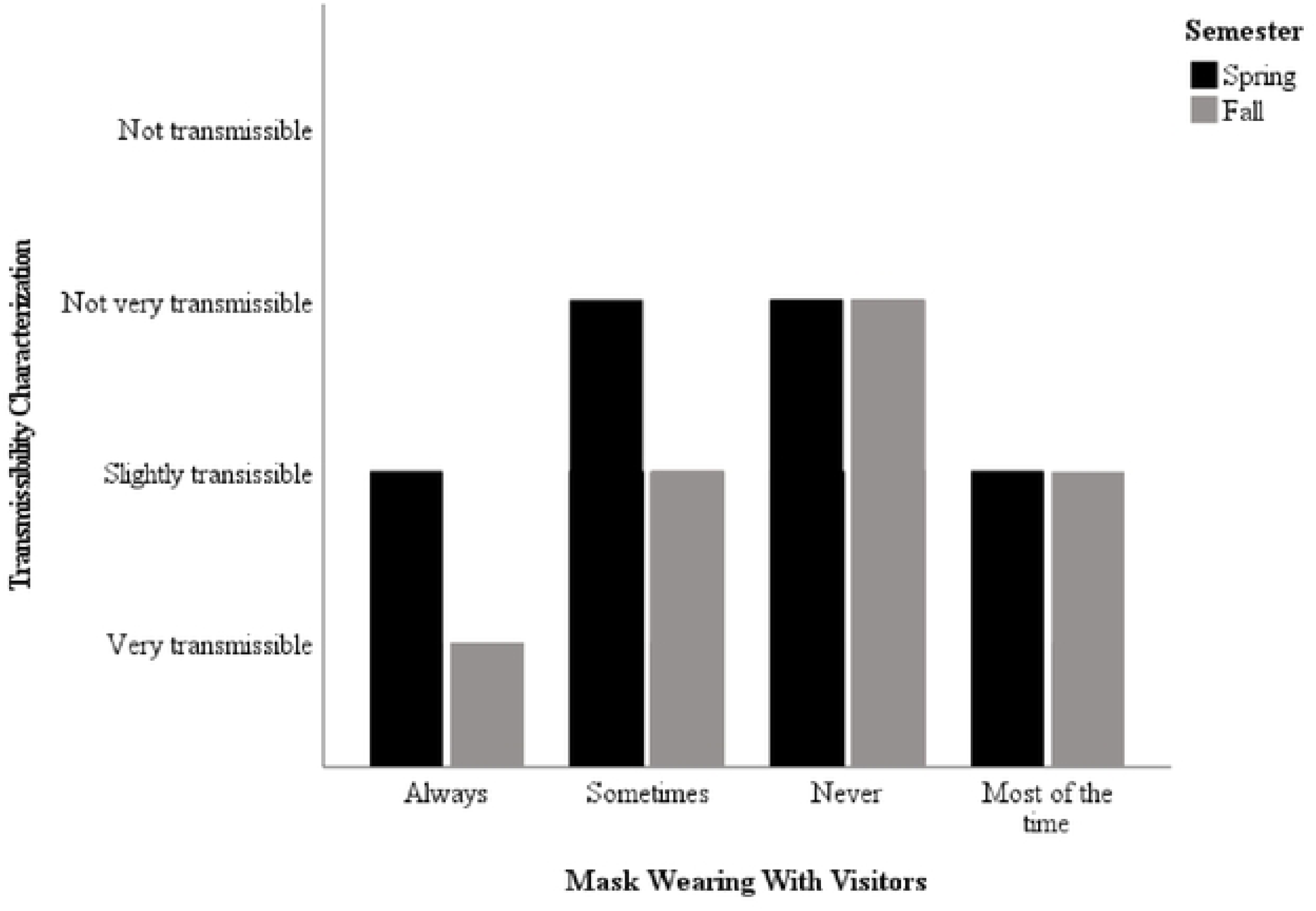
Reported Mask Wearing Among Students When Having Visitors Compared to their Perceptions on Virus Transmissibility by semester.

Large gatherings were often discouraged during the COVID-19 outbreak. Survey responses showed 33.5% of students gathered in groups (e.g., parties, study groups, group hangouts) of 5+ on a weekly basis. There was an increase of persons reporting daily and weekly gatherings of 5+ in the Fall semester relative to the Spring and the differences were statistically significant (*p* < 0.001). No significant differences were observed between genders (*p* = 0.220). When comparing responses to perception of virus transmission, the majority of those that felt the virus was not very transmissible reported gatherings of 5+ at least weekly. The majority of those who reported not congregating in groups in 5+ also responded as viewing the virus as very transmissible (70.7%) (Figure 2.).

**Figure 2.**
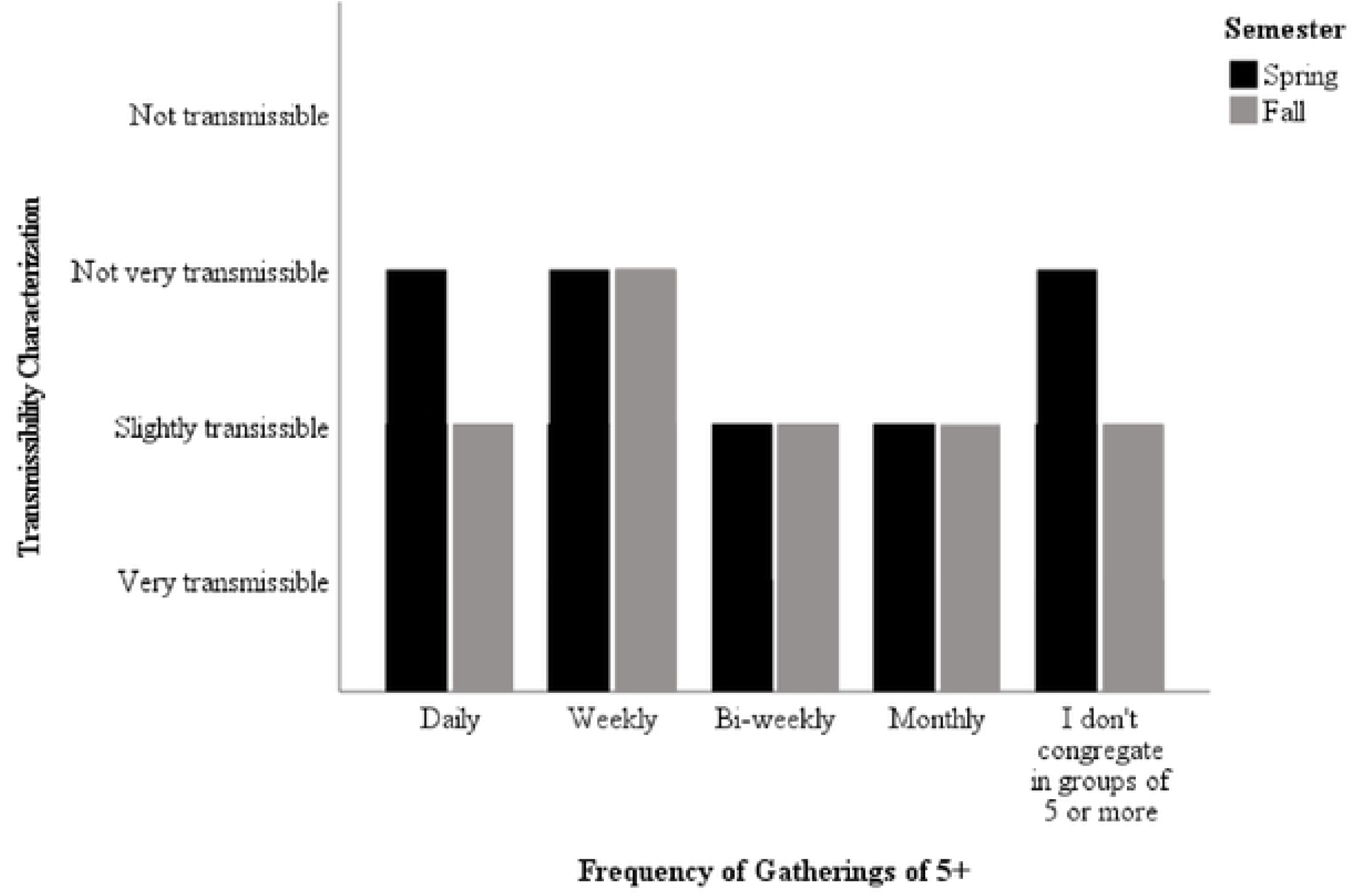
Reported Frequency of Group Gatherings Among Students Compared to their Perceptions on Virus Transmissibility by semester.

### Student Perceptions of ECU and COVID During Fall and Spring 2021 Semesters

The full return to campus may have been intimidating for some students. The University often worked to make sure COVID-19 information was disseminated and that students were able to access the information. Students were asked questions on whether they felt ECU gave them proper instructions to follow during the COVID-19 pandemic. The majority of students (87%) felt they received proper guidance and chi-square analysis showed the responses were between semesters were not significant (*p* = 0.157). One of the ways in which students were able to determine if it was okay to come to campus or class was by using a COVID Daily Screening Tool. Students were asked to use this checklist prior to coming to campus. Over half the students surveyed (52.5%) reported never using the daily screening tool, while only 11% reported always using it. There were no significant differences in tool usage when comparing Fall and Spring semesters (*p* = 0.601). The COVID daily screening tool was sent out via text message and email. Students were polled to find their preferred medium, with most students preferring texts (46.5%) followed by email (37.5%).

Universities were also tasked with ensuring that student learning was not greatly affected by shutdowns or transitions to remote learning during times of outbreak. Interestingly, 67% of ECU students felt their overall learning experience during COVID-19 was either greatly reduced or somewhat reduced (Figure 3). While most classes during the Spring semester were online, Fall saw the pivot to more traditional face-to-face learning for most classes; however, larger classes, 50+, continued utilization of online learning. Results showed that the differences between Fall and Spring semesters with regards to perceptions of learning experience were not significant (*p* = 0.780). Of note, most students who are recognized as upperclassmen reported a greatly reduced or somewhat reduced learning experience. Another objective was to determine whether students had difficulty transitioning from online learning to face-to-face during Fall semester. All students reported some difficulties in the transition, with 62.5% reporting the transition was extremely difficult.

**Figure 3.**
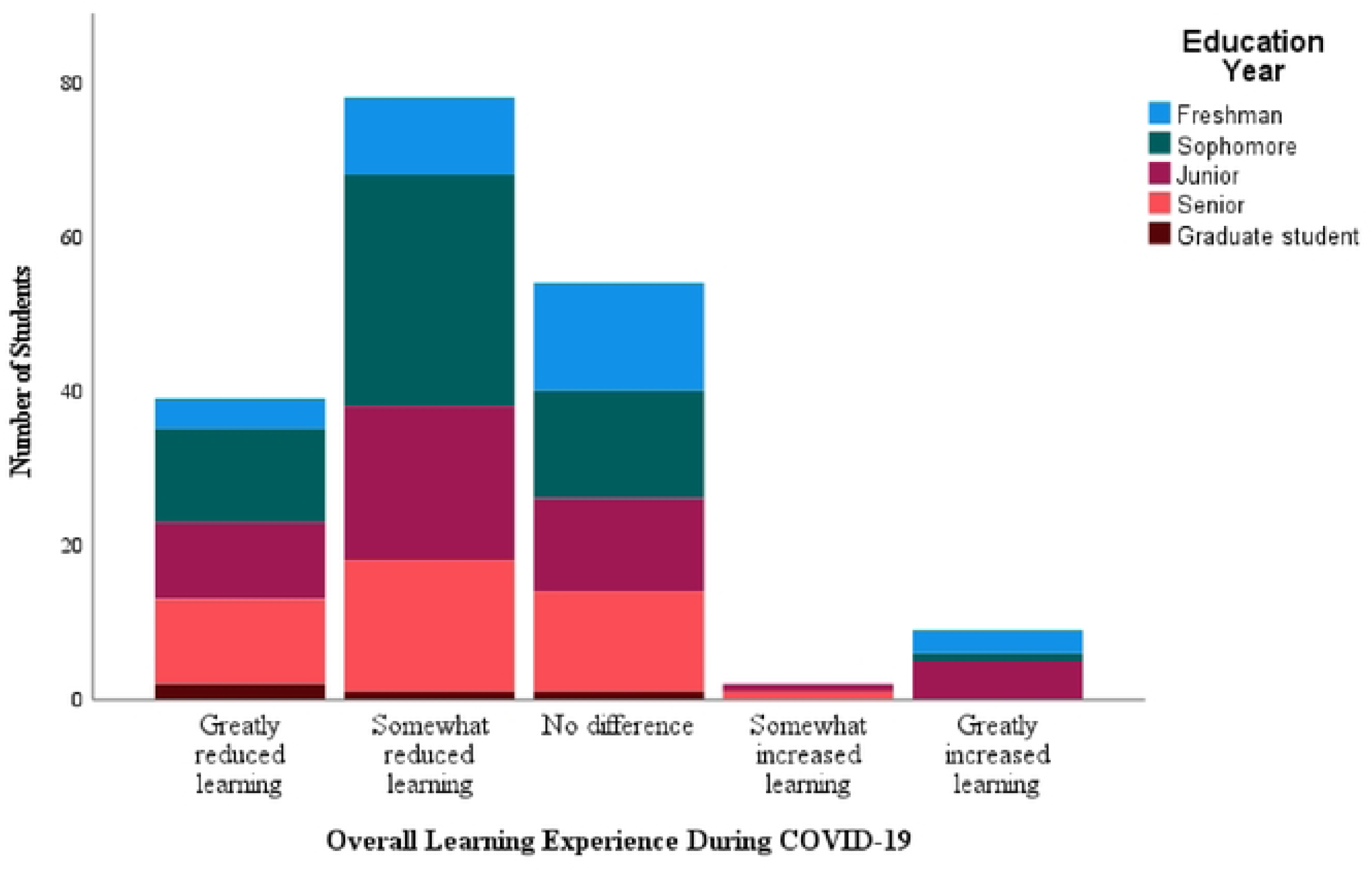
Overall learning experience during COVID-19 pandemic by education year.

Learning is a crucial part of student experience on college campuses, however social interactions and experience also play a role in student collegiate life. Overall, student responses showed most (80.8%) felt COVID-19 either greatly or somewhat affected their social activity. Students indicated that their overall social experience was better during the Fall relative to Spring semesters and the differences were statistically significant (*p* = 0.005), with 27.3% of Fall students compared to 50.6% of Spring semester students reporting “greatly reduced social activity”. Hence, students felt better socially (i.e., visiting, going out, being around people, etc.,) during the Fall semester.

## Discussion

Viral outbreaks often have effects on daily activities. Thus, routine activities may be influenced by perceptions and beliefs. Most students were concerned about the COVID-19 pandemic, while also feeling it to be very transmissible. The pandemic did not have a large overall effect on student’s perception of safety on campus as most students felt safe throughout both semesters. Overall, most students felt the vaccine had some level of safety and would take it. However, 30% of respondents during the Spring semester reported they felt the vaccine was “not safe and would not take it”. This may have been due to the origination of the vaccine during the Spring and the confusion over vaccine information (e.g., number of doses, potential side effects, etc.,). Additionally, students may have been concerned with the quickness in which the vaccine was developed, accepted, and dispersed to the general population. Indeed, a 2020 study by Lucia et al., highlighted a survey showing vaccine hesitancy among medical students.^22^ Another recent study on vaccine hesitancy highlighted vaccine acceptance rates of 77.6% for COVID-19 vaccine among general population along compared with an acceptance rate of 69% for the influenza vaccine among the general population.^24^ Interestingly, respondents noted some of the same reasons for hesitancy as students in our study such as safety, vaccine produced to quickly, and lack of trust. As universities sought to help students with these changes and perceived impacts on learning, there was an emphasis on reopening and resuming normal (face-to-face) classes. Part of this road to “normalcy” was an emphasis on students receiving vaccinations. Significant differences between semesters on the perceived safety of vaccines was observed. Students during the Fall semester had more positive perceptions and were more willing to take the vaccine. This may have been attributed to the fact that by the Fall semester, vaccination rollouts had been going on for numerous months and students were able to observe vaccine effects. Another reason that the perception of the vaccine may have changed could be that students were ready to return to campus. This change in perception and the addition of university vaccine encouragement may have aided in the University’s 74% achieved vaccination rate among on-campus students. As vaccines were heavily suggested, students may have felt they were safer. Universities, such as ECU, implemented tools such as COVID daily screening tools to help students determine the safety of coming to campus and/or to class. While these tools were disseminated via the preferred platforms of use by students: texts (46.5%) and emails (37.5%), over half of students surveyed reported never using the tool. Usage of the tool was low during each semester. It is possible that students did not use the tool because they felt they had been given proper guidance on protocols to take to help stem the virus. Students reported they felt ECU had given them proper guidance during the pandemic. Thus, it’s possible students felt that if they followed this guidance there was no reason to use the daily screening tool and that the tool was functioning as a redundancy.

Another objective was to determine if student behaviors differed between semesters as the pandemic continued. Behavioral guidance was issued to help stem virus spread which included mask wearing and discouraging large gatherings. Differences in mask wearing were observed between Spring and Fall semesters with more persons reporting never wearing a mask during the Fall semester when having visitors or when out visiting. Significant differences between semesters were observed as more students reported gathering in groups of 5+ persons during the Fall. This may have been attributed to multiple factors. By the Fall semester, vaccines were readily available and at the beginning of the semester many students, faculty, and staff had obtained vaccinations. Thus, students may have felt a sense of “herd immunity” and safer being in large groups. Additionally, many persons began to feel COVID burnout or fatigue as the pandemic continued on. Many people tired of restrictions and the feeling of helplessness.^25^ It has been observed many persons, particularly those in healthcare experienced fatigue and burnout.^26^ It is also possible students suffered from feelings of lost youth. Feelings such as these may have impacted behaviors, resulting in reduced caution among students.

### Learning Quality

The COVID-19 pandemic caused many schools and businesses to shut down and limit occupancy and activity in their buildings. Many campuses switched to online learning to help stem the spread of the virus while still allowing students to take needed classes. While this solution allowed students to continue their educational goals, it was important for universities to determine whether these changes had any effects on their student populations. Changes in educational delivery mechanisms attributed to COVID-19 may have affected students learning. Results from this survey showed 67% of students felt their learning was either somewhat or greatly reduced due to COVID-19. A survey conducted among 800 Polish medical students, showed while many felt online learning was enjoyable, they felt e-learning or online learning was statistically (*p* <0.001) less effective than traditional learning.^27^ This may be attributed to classes increasing in rigor and intensity. Additionally, students may have felt a disconnect with their instructor. It is possible not being a classroom may have impacted their ability to form a comfortable bond with their instructor, thus impacting their comfort level in asking follow-up or clarifying questions. Students here also reported difficulty in the transition to online learning. This falls in line with other studies that saw difficulties in transitioning to online platforms. One such study cited students as having increased stress and anxiety, difficulties concentrating along with technological and instructional challenges in the transition to online learning.^28^ These challenges may indeed have impacted students here, particularly technological as East Carolina University has a large population from rural areas. This appeared to be similar to students surveyed here, feeling they experienced a reduction in learning while using the online platform.

## Conclusion

Surveys may be instrumental in helping colleges and universities learn of student perceptions of important issues. This may have a profound effect on compliance with mitigation strategies such as vaccines, mask wearing, and social behavior adjustments. This knowledge of how students perceive the severity of a pandemic or public health emergency may also play a role in policy development aimed at helping to ensure safety and continuity of education for students. Information gathered from a representative sample of the population may inform public health officials of concerning behaviors that may exacerbate outbreaks and thus allow a more targeted approach. If universities are aware of student perceptions, they may be able to develop guidance based on these perceptions perhaps influencing students’ acceptance and compliance of mitigation protocols.

## Data Availability

Data has been provided as a part of this article.

## Acknowledgments

The authors would like to thank Will Bullock and East Carolina Student Affairs for their help in providing student housing data.

